# Amino acid profiling of COVID-19 patients blood serum

**DOI:** 10.1101/2024.03.05.24303773

**Authors:** Ya.V. Russkikh, N.N. Sushentseva, O.S. Popov, S.V. Apalko, V.S. Shimansky, A.Yu. Asinovskaya, S.V. Mosenko, A.M. Sarana, S.G. Scherbak

## Abstract

Main objectives of this study were to analyse metabolomic profile features of patients with COVID-19 using mass spectrometry techniques while taking into account the clinical and laboratory history, and to study the relationship between the severity of COVID-19 symptoms and the concentration of primary metabolites, primarily amino acids. We used frozen blood serum samples of 935 COVID-19 patients from the City Hospital No. 40 biobank collection. Metabolomic profile was studied by HPLS-MS/MS method. R programming language was used for statistical data processing. The difference of metabolic profile of patients with COVID-19 depending on the severity of the disease was revealed based on the performed analysis - for 52 out of 84 detected compounds there were differences with reliability p<0,01. Statistically significant differences in concentration were recorded for organic acids, amino acids and their derivatives. Using samples from the biobank collection, a metabolomic study of the biomaterial of patients hospitalised with the diagnosis of COVID-19 was carried out. According to the results obtained, kynurenine, phenylalanine and acetylcarnitine were associated with the severity of COVID-19 infection.

## INTRODUCTION

Recently, there has been an increasing number of studies devoted to the analysis of the blood metabolome for diagnosis and assessment of disease severity ^1–4^. Metabolomic profiling of human biological fluids is of great interest in terms of opportunities to obtain additional knowledge about the pathogenesis of diseases and possible therapeutic targets. Identification of factors that allow predicting the risk of severe disease and its complications is an important task.

The main method of metabolome research is mass spectrometry (MS), which can determin a wide variety of compounds, including disease markers, even at very low concentrations with high reliability, specificity and sensitivity. Amino acid spectrum reflects many processes occurring in the body and may contain information about the course of many diseases. Plasma amino acids link all organ systems and play an important physiological role as basic metabolites and regulators of metabolism. Accordingly, metabolic changes occurring in diseases of various systems and organs will affect amino acid profiles, and the amino acid spectrum may serve as a prognostic factor or indicate the development of some diseases ^5^.

To date, many papers about metabolomic profile of patients with COVID-19 have been published. Most of them are aimed at determining the unique metabolic signature of COVID-19 or at predicting the severity of the disease and/or mortality from it ^1–3,6^. It has been noted that in many diseases, changes in the levels of certain amino acids can serve as a prognostic factor ^4,7^.

Serum amino acid concentrations are quite strongly associated with COVID-19 symptomatology ^1,6–10^. Nevertheless, the available data about the involvement of individual amino acids in pathological processes vary considerably. Nevertheless, involvement of amino acid in pathological processes vary considerably according to available data. Thus, several studies reported that the level of phenylalanine concentration was positively associated with symptom severity ^7,11,12^, whereas other studies reported an inverse relationship ^13^ or no such relationship ^9,14^.

The purpose of this work was to identify the features of the metabolomic (primarily amino acid) profile of patients infected with COVID-19 with different severity and outcomes of the disease.

## RESULTS

Concentration data for 84 compounds with non-zero values in at least a few repetitions of measurements were used for analysis. The results of the primary metabolites analysis revealed differences in metabolomic profile according to disease severity for 52 out of 84 detected compounds (with significance p<0.01). The following metabolites showed the greatest difference when comparing groups by COVID-19 course severity: kynurenine (p<0.0001; lfc=1.62), phenylalanine (p<0.0001; lfc=1.13) and acetylcarnitine (p<0.0001; lfc=1.28).

Differences between groups with a high level of statistical significance were also found for the concentration of the following compounds: citrulline, ornithine, cystine, dimethylglycine, asymmetric dimethylarginine (ADMA), alanine, cystathionine, carnosine, γ-glutamylcysteine, arginine (Table).

In addition, tyrosine, histidine, proline, threonine, symmetric dimethylarginine (SDMA), creatinine, adenosine, thymidine monophosphate (TMP), cytosine, S-Adenosyl-L-homocysteine (SAH), and nicotinamide levels changed consistently with the progression of COVID-19 infection severity.

Differences in the metabolic profile between patients from group I and group II were observed for more than 30 compounds (Table). Elevated levels of kynurenine, phenylalanine, acetylcarnitine, dimethylglycine, proline, cytosine, tyrosine, adenosine, SAH, TMP, and to a lesser extent isoleucine, allantoin, asparagine, aspartate, 4-hydroxyproline, cystathionine were detected for group II patients, lysine, glycine, γ-glutamylcysteine, glutathione, valine, serine, adenosine monophosphate (AMP), α-ketoglutarate, malate, orotate, creatine, citrate, α-aminobutyrate, niacin, hypoxanthine, homocystine, choline, uridine, creatinine, carnitine, nicotinamide, ophthalmic acid, carnosine, guanosine, adenine, adrenaline, histamine. At the same time, the levels of threonine, ornithine, citrulline, cysteine, ADMA, SDMA, histidine, arginine, alanine, isocitrate, pyruvate, AMP, glycine, norepinephrine, S-adenosyl methionine (SAM), and cholate were lower in group II than in group I.

The metabolic profile of group III patients was characterized by further changes in the levels of the same compounds that distinguished group I from group II. Additionally, differences in the levels of isocitrate, AMP and uracil were noted.

Meanwhile, the metabolomic profiles of groups I and III differed in levels of 4-hydroxyproline, allantoin, SDMA, uridine, adenosine and SAH.

Differences in metabolomic profiles between surviving and deceased patients were observed for 37 compounds out of 82 (Table). Differences with a high level of significance were recorded for cystine (p<0.0005; lfc=1.36), cysteine (p<0.0001; lfc=1.81) and dimethylglycine (p<0.0001; lfc=1.29). Significantly lower levels of threonine, cysteine, homocystine, hydroxylysine, hypoxanthine, isocitrate, pyruvate, cholate, and serotonin were observed in the metabolome of deceased patients. At the same time, the levels of kynurenine, phenylalanine, citrulline, cytosine, dimethylglycine, histamine, γ-glutamylcysteine, cystine, carnitine, creatinine, creatine, cystathionine, leucine, isoleucine, AMP, TMF, SAH, acetylcarnitine, α-ketoglutarate, malate, lactate, urate, fumarate, nicotinamide, pantothenic acid and dopamine were higher than in surviving patients.

DISCUSSION

According to the results obtained, kynurenine, phenylalanine and acetylcarnitine have strong association with the severity of COVID-19 infection. This is consistent with the findings of increased levels of kynurenine and phenylalanine for COVID-19 patients ^3,8,15,16^ and their negative correlation with the severity of infection ^7,10^. Increased kynurenine levels in COVID-19 are associated with increased tryptophan degradation due to overactivation of the immune response through increased levels of interferon-gamma (increased inflammatory response) and strong T-cell activation ^1,15^; kynurenine pathway metabolites were shown to be associated with tricarboxylic acid cycle intermediates, inflammatory response, and cell death ^4^.

Acetylcarnitine plays an essential role in energy metabolism and transport of fatty acids into mitochondria. An imbalance of acetylcarnitine in COVID-19 infection has been reported in a number of studies ^3,11,15^.

A strong correlation with COVID-19 infection severity was also recorded for citrulline, ornithine, cystine, dimethylglycine, ADMA, alanine, cystathionine, carnosine, γ-glutamylcysteine, and arginine.

In addition, levels of tyrosine, adenosine, histidine, creatinine, TMP, proline, threonine, cytosine, SDMA, SAH, nicotinamide were consistently altered as the severity of COVID-19 infection progressed, which is consistent with the data on the correlation of tyrosine biosynthesis pathways with the severity of COVID-19 infection ^17^[17], as well as the relationship between the severe course of COVID-19 and levels of such amino acids as phenylalanine, proline, valine, valine, glutamate, glutamine, tryptophan, histidine, alanine, leucine, isoleucine, cysteine ^18,19^.

Impaired synthesis and metabolism of arginine, threonine, ornithine, citrulline and alanine in COVID-19 patients, especially under hypoxic conditions, has been reported in many studies ^9,11,16,19^. Decreased serum levels of glutamate, citrulline, ornithine, glutamine, urea, fumarate, ADMA and SDMA in COVID-19 patients were associated with liver dysfunction ^14^.

Metabolic profile of patients with COVID-19 in severe condition (group III) was characterized by significantly greater changes compared with groups I and II (Table 3) and affected all classes of metabolites studied, indicating systemic metabolic disorders resulting from the development of COVID-19 and affecting the functioning of various organs and systems of the body.

**Table 1:** Sex and age structure of the examined groups of patients [*M ± SD*].

| Comparison | Group | n | n (M) | n (F) | Age | Age (M) | Age(F) |
| --- | --- | --- | --- | --- | --- | --- | --- |
| Disease course severity (n=935) | I | 25 | 16 | 9 | 51 ± 16, 2 | 50, 9 ± 16, 6 | 51, 3 ± 16, 3 |
|  | II | 564 | 263 | 301 | 61, 5 ± 15, 5 | 59, 7 ± 15, 3 | 63, 1 ± 15, 5 |
|  | III | 346 | 166 | 180 | 69, 1 ± 14, 2 | 67, 1 ± 14, 3 | 70, 9 ± 13, 8 |
| Disease outcome (n=935) | IV | 141 | 73 | 68 | 74, 8 ± 12, 4 | 73 ± 12, 1 | 76, 7 ± 12, 5 |
|  | V | 794 | 372 | 422 | 62, 1 ± 15, 3 | 60 ± 15, 2 | 64 ± 15, 2 |

Note: I - group of patients with mild course of the disease, II - group of patients with moderately severe course of the disease, III - group of patients with severe course of the disease, IV - group of patients with fatal outcome of the disease, V - group of patients successfully discharged after hospitalisation. Comparison - parameter by which the studied groups are compared, in brackets [n] is given the total number of patients from the sample included in the groups. M - male, F - female.

**Table 2:** List of primary metabolites.

|  |  |
| --- | --- |
| **Glycolysis** | **Organic acids** |
| Lactic acid | γ-Aminobutyric acid |
| Pyruvic acid | Adenylosuccinate |
| **Citric acid cycle** | Argininosuccinic acid |
| α-Ketoglutaric acid | Cholic acid |
| Aconitic acid | Creatine |
| Citric acid | Niacin |
| Fumaric acid | Ophthalmic acid |
| Isocitric acid | Orotic acid |
| Malic acid | Pantothenic acid |
| Succinic acid | Taurocholic acid |
| **Amino acids** | Uric acid |
| 4-Hydroxyproline | **Nucleosides and nucleotides** |
| Alanine | Adenine |
| Arginine | Cytosine |
| Asparagine | Guanine |
| Aspartic acid | Thymine |
| Lysine | Uracil |
| itrulline | Xanthine |
| Cystine | Adenosine |
| Dimethylglycine | Cytidine |
| Glutamic acid | Guanosine |
| Glutamine | Inosine |
| Glycine | Thymidine |
| Histidine | Uridine |
| Homocystine | 3',5'-cyclic adenosine monophosphate |
| Isoleucine | AMP |
| Leucine | 3',5'-cyclic cytidine monophosphate |
| Asymmetric dimethylarginine (ADMA) | Cytidine monophosphate |
| Methionine sulfoxide | 3',5'-cyclic guanosine monophosphate |
| Symmetric dimethylarginine (SDMA) | Guanosine monophosphate |
| Phenylalanine | Thymidine |
| Proline | **Other compounds** |
| Serine | α-Aminobutyric acid |
| Ornithine | Acetylcarnitine |
| Threonine | Acetylcholine |
| Tryptophan | Allantoin |
| Tyrosine | Carnitine |
| Valine | Carnosine |
|  | Choline |
| **Transsulfuration pathway** | Citicoline |
| Cystathionine | Creatinine |
| Cysteine | Cysteamine |
| Homocysteine | 3,4-dihydroxyphenylalanine |
| Methionine | Dopamine |
| γ-glutamylcysteine | Adrenaline |
| Glutathione | Histamine |
| Oxidized glutathione | Hypoxanthine |
| S-adenosyl-L-homocysteine (SAH) | Kynurenine |
| S-adenosyl methionine (SAM) | Nicotinamide |
|  | Norepinephrine |
| **Cofactors** | Serotonin |
| FAD (Flavin adenine dinucleotide) | **Internal standards** |
| FMN (Flavin mononucleotide) | 2-(N-morpholino)ethanesulfonic acid |
| NAD (Nicotinamide adenine dinucleotide) | L-Methionine sulfone |

**Table 3:**
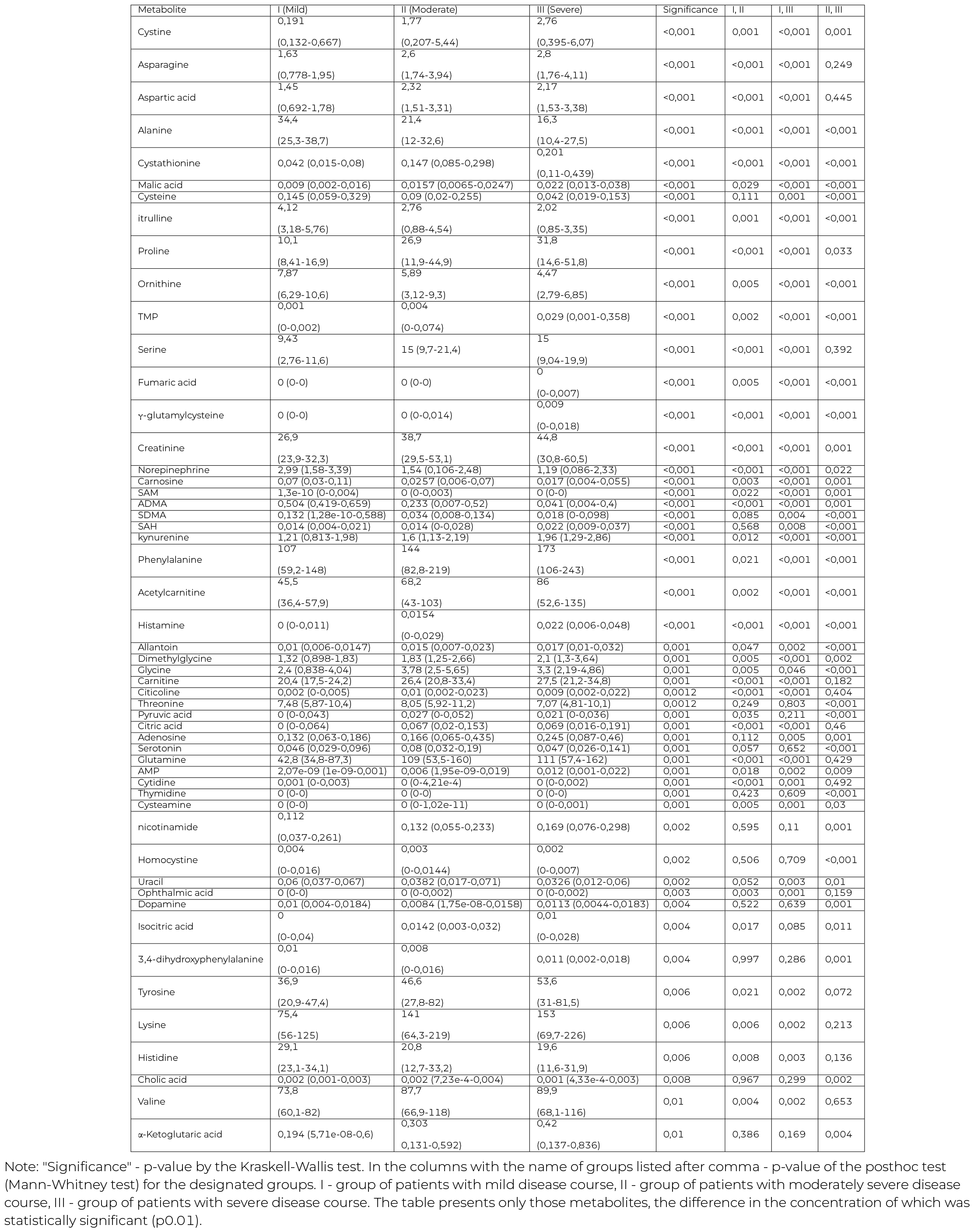
Metabolite concentrations in the blood of patients from different groups divided by disease severity [Me (Q25-Q75)], numbers less than 0.001 are presented by exponential entry [number e degree].

**Table 4:** Metabolite concentrations in the blood of patients from different groups divided by disease outcome [Me (Q25-Q75)], numbers less than 0.001 are presented by exponential entry [number e degree].

| Metabolite | V (Alive) | IV (Dead) | p |
| --- | --- | --- | --- |
| Threonine | 7,88 (5,71-11,1) | 6,9 (4,32-8,93) | <0,001 |
| Cystathionine | 0,15 (0,085-0,303) | 0,251 (0,126-0,769) | <0,001 |
| Malic acid | 0,017 (0,008-0,026) | 0,023 (0,013-0,045) | <0,001 |
| Isocitric acid | 0,014 (0,002-0,032) | 0,005 (0-0,022) | <0,001 |
| Pyruvic acid | 0,026 (0-0,049) | 0,016 (0-0,031) | <0,001 |
| Lactic acid | 12,3 (8,71-17,1) | 15,6 (11,3-22,9) | <0,001 |
| Uric acid | 3,91 (2,7-5,66) | 5,17 (3,46-7,85) | <0,001 |
| Creatine | 13 (7,94-21,9) | 17,1 (9,61-40,3) | <0,001 |
| Creatinine | 38,9 (29,2-53,1) | 49,3 (32,7-70,5) | <0,001 |
| Pantothenic acid | 0,089 (0,063-0,118) | 0,106 (0,077-0,156) | <0,001 |
| Dopamine | 0,009 (0,001-0,016) | 0,0135 (0,006-0,019) | <0,001 |
| SAH | 0,015 (0-0,028) | 0,032 (0,018-0,055) | <0,001 |
| kynurenine | 1,63 (1,14-2,26) | 2,4 (1,5-3,5) | <0,0001 |
| phenylalanine | 145 (83,9-218) | 202 (143-258) | <0,0001 |
| acetylcarnitine | 68,1 (43-104) | 110 (68,1-175) | <0,001 |
| 3,4-dihydroxyphenylalanine | 0,008 (0-0,016) | 0,013 (0,006-0,019) | <0,001 |
| $\alpha$ – *Ketoglutaricacid* | 0,307 (0,129-0,598) | 0,581 (0,149-1,09) | 0,001 |
| TMP | 0,006 (0-0,104) | 0,025 (0,002-0,348) | 0,001 |
| Homocystine | 0,003 (0-0,014) | 0,001 (0-0,004) | 0,001 |
| citrulline | 2,63 (0,89-4,29) | 1,97 (0,78-3,04) | 0,001 |
| Hydroxylysine | 0 (0-0,136) | 0 (0-0,009) | 0,001 |
| Histamine | 0,0162 (0-0,031) | 0,025 (0-0,056) | 0,001 |
| cystine | 1,88 (0,208-5,49) | 2,83 (1,37-6,11) | 0,001 |
| AMP | 0,006 (1,58e-09-0,012) | 0,014 (0,001-0,024) | 0,001 |
| Fumaric acid | 0 (0-2,11e-08) | 0 (0-0,009) | 0,001 |
| Lysteine | 0,0749 (0,021-0,241) | 0,0344 (0,0194-0,123) | 0,001 |
| Cytosine | 0,013 (0,003-0,028) | 0,021 (0,004-0,046) | 0,001 |
| Hypoxanthine | 0,786 (0,387-1,29) | 1,07 (0,504-1,72) | 0,001 |
| carnitine | 26 (20,4-32,9) | 28,6 (23,1-39,7) | 0,001 |
| Cholic acid | 0,002 (6,61e-4-0,004) | 9,92e-4 (0,0002-0,003) | 0,001 |
| Thymidine | 0 (0-0) | 0 (0-0) | 0,001 |
| Isoleucine | 43 (27,2-74,4) | 56,4 (34,6-82,6) | 0,003 |
| Serotonin | 0,0695 (0,03-0,181) | 0,0389 (0,025-0,123) | 0,003 |
| $\gamma$ – *glutamylcysteine* | 0 (0-0,015) | 0,01 (0-0,017) | 0,003 |
| Nicotinamide | 0,136 (0,058-0,251) | 0,184 (0,089-0,286) | 0,003 |
| Leucine | 67,3 (41,7-110) | 87,7 (55,5-123) | 0,009 |
| dimethylglycine | 1,83 (1,25-2,72) | 2,28 (1,34-4,1) | 0,0099 |

Note: IV - group of patients with fatal outcome of the disease, V - group of patients successfully discharged after hospitalisation. "Significance" - p-value by the Mann-Whitney test. The table presents only those metabolites, the difference in the concentration of which is statistically significant (p0.01).

Citrullin is produced almost exclusively by enterocytes and is used as a biomarker of small intestinal enterocyte mass and function. COVID-19 can infect human intestinal cells and also replicate in intestinal epithelial cell lines and organoid models of human colon, thus participating in the spread of COVID-19 with increased viremia ^20^. A study of differences in plasma amino acid levels between the acute and convalescent stages in people with community-acquired pneumonia showed that plasma levels of arginine and citrulline decreased ^19^, which is consistent with our findings.

Elevated plasma cytosine levels in COVID-19 patients may be associated with virus escape from innate immunity ^21^, and cytosine-based metabolites are coordinators of cellular metabolism in COVID-19, and are important for innate antiviral immunity and virus evolution. Observed significant increase of creatine and creatinine in metabolomic profiles of deceased patients compared to survivors indicates the development of renal dysfunction ^15^.

The most significant differences in metabolic profiles between surviving and deceased patients were recorded for cystine, cysteine and dimethylglycine. Reduction of dimethylglycine level in patients with unfavorable course of COVID-19 was also noted by Silvagno F, et al. ^22^.

Decreased glutathione levels were observed ^22,23^ in diseases that increase the risk of COVID-19, and the resulting glutathione deficiency was associated with the severity of course and death in patients with COVID-19. An increase in cystine and cysteine levels in the plasma of COVID-19 patients with increasing severity of infection (from moderate to severe) was also reported in Bramer et al. ^24^. Cystine is formed in the extracellular space from reduced glutathione, functions as a general marker of infection and can serve as an indicator of glutathione production and activity of its cycle under oxidative stress ^22,24^.

Imbalance of cystine and cysteine depending on COVID-19 disease severity, immune activity and presence of comorbidities was also reported by Páez-Franco JC ^25^[25]. It has been observed that cystine levels are crucial for the control of reactive oxygen species in COVID-19 as well as in some malignancies. Alanine transaminase, aspartate transaminase and cystine levels correlation in severe COVID-19 patients may indicate a potential involvement of the liver in cysteine and cystine metabolism in COVID-19, which may be likely since this organ is the main site of glutathione synthesis and often is affected in severe cases of COVID-19^25^.

Cysteine/cystine, cysteinylglycine/cystinylglycine and glutathione/glutathione disulfide are the main redox pairs for maintaining of extracellular thiol-disulfide balance. They are indicators of an age-related decrease in systemic reduction potential and the influence of this process on various tissues, including the pulmonary ^26^. COVID-19 disrupts cellular the cystine/cysteinic cycle and extracellular thyols redox homeostasis mechanisms. It promotes the replication of the virus due to the formation of pro-oxidants in the infected tissue. Indeed, the preferential incorporation of cellular cysteine into viral proteins rather than into glutathione of cellular proteins has been observed as a common mechanism in other types of viral infections ^26^.

Increased SAM levels in the metabolomic profiles of patients with high comorbidity index values can be considered as a marker of lung damage risk in COVID-19 patients and possibly as a factor associated with the development of inflammation, since SAM and SAH are indicators of transmethylation and may play an important role as markers of COVID-19 severity ^27^.

## MATERIALS AND METHODS

### Participant Characteristics

A frozen blood serum from the City Hospital No 40 biobank collection was used to amino acids profiling. The study was approved by the expert ethics board of the St. Petersburg State Health Care Institution “City Hospital No. 40” (protocol No. 171 dated May 18, 2020). All donors had signed voluntary informed consent to participate in the study.

Serum samples from 935 patients (445 males and 490 females) were included in the study (Table 1)

Participation criteria:

1. Positive PCR test for COVID-19
2. Voluntary informed consent to participate in the study.
3. No history of HIV, hepatitis B and C, or syphilis. 4.
4. Blood collection was performed before anticytokine therapy, hemosorption, transfusion of blood and its components.

Patients were divided into groups according to the severity of the disease. Severity level was determined by the current version of the “Interim Guidelines. Prevention, diagnosis and treatment of novel coronavirus infection (COVID-19)”: (I) “mild” - group of patients with mild COVID-19 infection without changes on a computed tomography (CT) scan characteristic of a viral lesion (16 males and 9 females); (II) - “moderate” - group of patients with elevated body temperature (>38 degrees C), relatively low blood oxygen saturation and changes on a CT scan typical of a viral lesion (263 men and 301 women), (III) - “severe” - group of patients with a severe course of COVID-19 infection, with decreased level of consciousness, unstable hemodynamics and changes on CT scan up to critical degree of lesion and development of acute respiratory distress syndrome (166 men and 180 women). Group IV – patients with fatal outcome of the disease (total 73 men and 68 women), and surviving patients - group V - were used as a comparison group.

### Metabolite profile study method

2-(N-morpholino)ethanesulfonic acid and L-methionine sulfone (Sigma-Aldrich) were used as internal standards. The samples were thawed at room temperature, then 100 tl of a solution of the internal standard mixture in acetonitrile was added to 50 tl of serum. The obtained extract was diluted with water. The prepared standard solutions and extracts were stored at -20 C.

The metabolite profile was analyzed using LCMS-8050 triple quadrupole liquid chromatography-mass spectrometer (Shimadzu) with Nexera X2 chromatography system. HPLC-MS/MS - Analysis was performed using the commercially available “LC/MS/MS Method Package for Primary Metabolites” method for the analysis of primary metabolites using a Discovery HS F5-3 analytical column (150Œ2.1 mm, 3 m) (Supelco, Merck) in multiple reaction monitoring mode. The method allows simultaneous analysis of 98 primary metabo-lites (Table 2) including amino acids, organic acids, nucleotides, nucleosides, and coenzymes. Mass spectrometric and chromatographic conditions and parameters were set according to the method instruction “LC/MS/MS Method Package for Primary Metabolites”. Data was collected and processed using LabSolutions software according to the internal standard method.

### Statistical processing of data

The Shapiro-Wilk test was used to test the hypothesis of normal distribution of data. One-factor analysis of variance using the Kraskell-Wallis test was used to detect intergroup differences in the concentration levels of the compounds under study, and the Mann-Whitney test was used as a post-hoc analysis. The logarithm of fold change (lfc) was used as a measure of the difference in the range of values between samples, descriptive statistics was represented by the median (Me) and interquartile range [Q25-Q75]. Data processing and statistical analysis were performed using the R programming language version 4.3.1.

## Data Availability

All data produced in the present study are available upon reasonable request to the authors

## ACKNOWLEDGMENTS

The authors are grateful to the study participants and the staff from the St. Petersburg State Health Care Institution City Hospital No. 40, Kurortny District hospital. The authors would like to thank all authors of the included studies for their valuable contributions to data collection, and Saint-Petersburg University R&D grant for making this study possible.

